# Plasma Glial Fibrillary Acidic Protein and Neurofilament Light are Elevated in Bipolar Disorder: Evidence for Neuroprogression and Astrocytic Activation

**DOI:** 10.1101/2024.07.30.24311203

**Authors:** Matthew Kang, Dhamidhu Eratne, Olivia Dean, Michael Berk, Adam J Walker, Cassandra Wannan, Charles B Malpas, Claudia Cicognola, Shorena Janelidze, Oskar Hansson, Jasleen Grewal, Philip B Mitchell, Malcolm Hopwood, Christos Pantelis, Alexander F Santillo, Dennis Velakoulis MiND Study Group

**Affiliations:** Neuropsychiatry Centre, Royal Melbourne Hospital; Department of Psychiatry, University of Melbourne; Deakin University and Barwon Health, Institute for Mental and Physical Health and Clinical Translation (IMPACT), School of Medicine, Geelong, Victoria, Australia; Florey Institute for Neuroscience and Mental Health, Parkville, Victoria, Australia; Deakin University and Barwon Health, Institute for Mental and Physical Health and Clinical Translation (IMPACT), School of Medicine, Geelong, Victoria, Australia; Orygen, Parkville, Victoria, Australia; Centre for Youth Mental Health, The University of Melbourne, Parkville, Victoria, Australia; Department of Medicine, Melbourne Medical School, University of Melbourne; Melbourne School of Psychological Sciences, University of Melbourne; Clinical Memory Research Unit, Department of Clinical Sciences Malmö, Faculty of Medicine, Lund University, Lund, Sweden Memory Clinic, Skåne University Hospital, Malmö, Sweden; Clinical Memory Research Unit, Department of Clinical Sciences Malmö, Lund University, Lund, Sweden; Alfred Mental and Addiction Health, Alfred Health, Melbourne, Victoria, Australia; Discipline of Psychiatry and Mental Health, School of Clinical Medicine, Faculty of Medicine and Health, UNSW; Department of Psychiatry, University of Melbourne Professorial Psychiatry Unit, Ramsay Clinic Albert Road; Department of Psychiatry, The University of Melbourne, Parkville, VIC, Australia, Western Centre for Health Research & Education, University of Melbourne & Western Health, Sunshine Hospital, St Albans, Victoria, Australia, Monash Institute of Pharmaceutical Sciences (MIPS), Monash University, Parkville, Melbourne, Victoria, Australia; Department of Clinical Sciences, Clinical Memory Research Unit, Faculty of Medicine, Lund University, Malmö, Sweden; Neuropsychiatry Centre, Royal Melbourne Hospital; Melbourne Neuropsychiatry Centre & Department of Psychiatry, University of Melbourne

**Keywords:** Biological markers, Bipolar Disorder, Neurofilament Light Chain, Glial Fibrillary Acidic Protein, Mania, Depression, Mental Health, Psychiatry

## Abstract

**Importance:** Recent methodological developments allow us to measure small amounts of brain-specific proteins in the blood, including neurofilament light chain (NfL), a marker of axonal pathology, and glial fibrillary acidic protein (GFAP), a marker of astrocytic activation. Given the evidence of potential astroglial pathology and neuronal dysfunction in bipolar disorder, these markers may provide further insight into its pathophysiology.

**Objective:** We investigated plasma NfL and GFAP levels in people with bipolar depression and compared them with unaffected individuals.

**Design, Setting, and Participants:** This cross-sectional study included 216 individuals, of which 120 participants had bipolar depression and 96 healthy controls. The blood samples were analysed between November 2023 and April 2024.

**Main outcomes and measures:** We used bootstrapped general linear models (GLM) to compare plasma NfL and GFAP levels between people with bipolar depression and healthy controls, adjusted adjusting for age, sex, and weight. We examined associations between these biomarkers and clinical variables, including mood symptom severity, past psychiatric history, and functioning, adjusting for multiple comparisons. For additional sensitivity analyses, predictors were evaluated using Bayesian model averaging (BMA).

**Results:** GFAP and NfL levels in plasma were elevated in people with bipolar depression (n = 120) compared to healthy controls (n = 96) after adjusting for age, sex and weight. The duration of illness was positively associated with NfL. The BMA analysis also identified duration of illness as a strong predictor of NfL (Posterior Inclusion Probability, PIP = 0.85). Age of onset was positively associated with GFAP. The BMA analysis similarly found age of onset to be a moderately strong predictor (PIP = 0.67).

**Conclusions and Relevance:** This study found elevated levels of plasma NfL and GFAP in bipolar depression compared to unaffected individuals, with significant associations with the duration of illness and age at onset, suggesting a degree of neuronal injury and astrocytic dysfunction in bipolar depression. These biomarkers may reflect specific illness stages, including neuroprogression and the later onset of bipolar disorder.

**Key Points:** Question: Is there neuronal dysfunction and astroglial pathology in bipolar disorder?

Findings: In this study, plasma levels of neurofilament light chain (NfL) and glial fibrillary acidic protein (GFAP) were elevated in individuals with bipolar disorder compared to unaffected individuals. These biomarkers correlated with the duration of illness and age of onset, indicating neuronal injury and astrocytic activation.

Meaning: These biomarkers may identify different stages and phenotypes of bipolar disorder based on their neurobiological underpinnings, providing potential diagnostic and prognostic value.

## INTRODUCTION

Bipolar disorder is a chronic psychiatric disorder characterised by periods of depression and mania/hypomania.^1^ Depressive phases account for the majority of time in bipolar disorder,^2,3^ leading to significant impairments in daily functioning and quality of life.^1^ Despite advances in understanding the clinical manifestations of bipolar disorder, the neurobiological underpinnings remain unclear, hindering the development of targeted treatments.^4^

Blood-based biomarker research investigating the pathophysiology of bipolar disorder has focused on systemic markers such as cytokines, which are not brain-specific.^4^ Recent technological advancements have made it possible to accurately measure picograms of brain-derived proteins in the blood.^5^ Two markers have drawn attention in the neurodegenerative disease space. Neurofilament light chain (NfL) is a cytoskeletal intermediate filament protein expressed in neurons, with elevated concentrations in blood following neuronal injury and degeneration.^6^ Glial fibrillary acidic protein (GFAP) is a component of the astrocyte cytoskeleton, with elevated concentrations being associated with astrocyte activation, a marker of neuroinflammation.^7^ These proteins are of particular interest in bipolar disorder, given that the condition has been linked with neuronal dysfunction, neuroinflammation.^8^ Additionally, the association of bipolar disorder with cognitive and functional decline has led to the “neuroprogression” hypothesis, suggesting that there is pathological rewiring of the brain over the course of the illness.^8–10^ By examining these brain-specific biomarkers, we can gain valuable insights into the neurobiological mechanisms underlying bipolar disorder.

Our recent systematic review^11^ found that there was mixed and limited literature as to whether NfL levels are elevated in bipolar disorder compared to healthy controls. The mood states of these cohorts at the time of blood sampling were varied (euthymic, depressed, or mixed). Two studies specifically explored bipolar depression and found that plasma NfL was mildly elevated compared to healthy controls.^12,13^ Only one study investigated GFAP in a small number of people with bipolar disorder found that serum GFAP levels were similar to healthy controls.^7^ None of these studies examined the correlation between plasma NfL/GFAP and clinical variables such as symptom severity.

Investigating NfL and GFAP in the blood of individuals with bipolar depression could provide valuable insights into the extent of neuronal pathology and astrocyte activation associated with the disorder, providing new avenues for therapeutic strategies. In this study, we aimed to further investigate the bipolar depression group by analysing plasma GFAP concentrations and comparing them with healthy controls. Furthermore, we aimed to build on our previous finding that NfL was higher in people with bipolar disorder,^13^ by exploring the relationships between NfL and GFAP with clinical and demographic factors in bipolar disorder to better understand what illness-related factors are associated with higher biomarker levels.

## METHODS

### Study Cohorts

This study followed the Strengthening the Reporting of Observational Studies in Epidemiology (STROBE) reporting guidelines,^14^ and included 210 individuals from two multicentre cohorts. The bipolar depression cohort were from a registered clinical trial (ACTRN12612000830897) of adjunctive mitochondrial agents and N-acetylcysteine.^15^ Participants were at least 18 years old, met DSM-IV-TR diagnostic criteria for bipolar disorder, and experiencing a bipolar depressive episode of at least moderate severity. Participants were required to remain on stable treatment for at least one month prior to the study.

The healthy controls were pooled from the Cooperative Research Centre (CRC) for Mental Health Study^16^ and had no current or past psychiatric or neurological illness. We have reported plasma NfL values for this cohort previously.^17^

### Ethics

This study, which is part of The Markers in Neuropsychiatric Disorders Study (The MiND Study, https://themindstudy.org), was approved by the Melbourne Health Human Research Ethics Committee (MH HREC 2020.142).

### Sample analysis

Plasma aliquots from all samples were stored at −80°C. Samples were randomised before analysis, and analyses were blinded to diagnosis. Plasma NfL and GFAP levels were measured on the Quanterix SR-X analyser using Simoa NF-Light kits and Simoa GFAP Discovery kits, according to the manufacturer’s guidelines (Quanterix Corporation, Billerica, MA USA). Both NfL and GFAP are suitable for retrospective analysis, as it is robust to multiple freeze-thaw cycles and time.^18–20^

### Statistical analysis

All statistical analyses were performed using *R* (Version 4.2.2).^21^ To compare the demographic variables between bipolar depression and healthy controls, we used Mann–Whitney U-tests and Pearson’s chi-square tests of independence.

General linear models (GLM) were used to compare NfL and GFAP levels between healthy controls and individuals with bipolar disorder. NfL and GFAP values were entered as dependent variables and were log_10_ transformed due to positively skewed distributions, and we reported raw values for easier interpretation. Cohort group, age, sex, and weight were entered as independent variables, and 95% confidence intervals were computed (nonparametric bootstrapping, 1000 replicates), with statistical significance defined as any confidence interval not including the null (at 95% level).

#### Exploratory analysis of clinicodemographic variables and biomarker values

##### Conventional frequentist analysis

For exploratory analysis of clinical and demographic factors that may explain the elevated biomarker level in bipolar disorder, we used Chi-squared tests of independence, Welch’s t-test and ANOVA correlation coefficients to analyses their associations. We adjusted for multiple comparisons using Benjamini-Hochberg false discovery rate (FDR) correction. All statistical assumptions were checked, including multicollinearity in linear models using the variance inflation factor (VIF).

##### Bayesian model averaging

To further mitigate the problem of using multiple predictors, we used Bayesian model averaging (BMA),^22^ a validated statistical technique that allows for all potential predictors to be examined in the model in parallel, while simultaneously considering all possible formulations of the model. The overall statistical solution is composed of all possible combinations of predictors weighted by model fit. As the best-fitting combinations contribute more to the final model, and the worst-fitting models contribute less, this naturally deals with the problem of model selection without having to impose model selection bias.

Briefly, for *p* predictors, BMA estimates all possible models corresponding to the 2^p^ possible combinations of predictors. The fit of each model was then evaluated using the log of the posterior odds. Parameter estimates were averaged over all models, weighted for the fit of each model, which inherently adjusted for the uncertainty associated with model selection bias.

The importance of each predictor was evaluated by the posterior inclusion probability (PIP) value, which was the probability that the parameter is not zero given the data. Predictors with PIP values > 0.5 were considered ‘important’. Predictions of NfL or GFAP for each patient were computed using the posterior predictive distribution averaged across all models. BMA was performed using the Bayesian adaptive sampling package.^23^ As one variable (number of depressive episodes) had 11 missing values, all BMA analyses were run with and without this variable to ensure that the results were consistent.

## RESULTS

### Study cohort characteristics

A total of 216 participants had adequate plasma samples available for NfL and GFAP analyses. The 120 participants with bipolar disorder comprised of 75 (62%) females and 45 (38%) males, with a mean ± SD age of 43.9 ± 12.0 years and weight of 84.3 ± 21.0kg. For healthy controls (n=96), there were 50 (52%) females and 46 (48%) males, with a mean ± SD age of 44.7 ± 14.4 years and weight of 77.3 ± 15.4kg. The bipolar depression cohort and the healthy controls were not statistically different in terms of their demographic measures, including their weight (**Table 1**).

**Table 1-.**
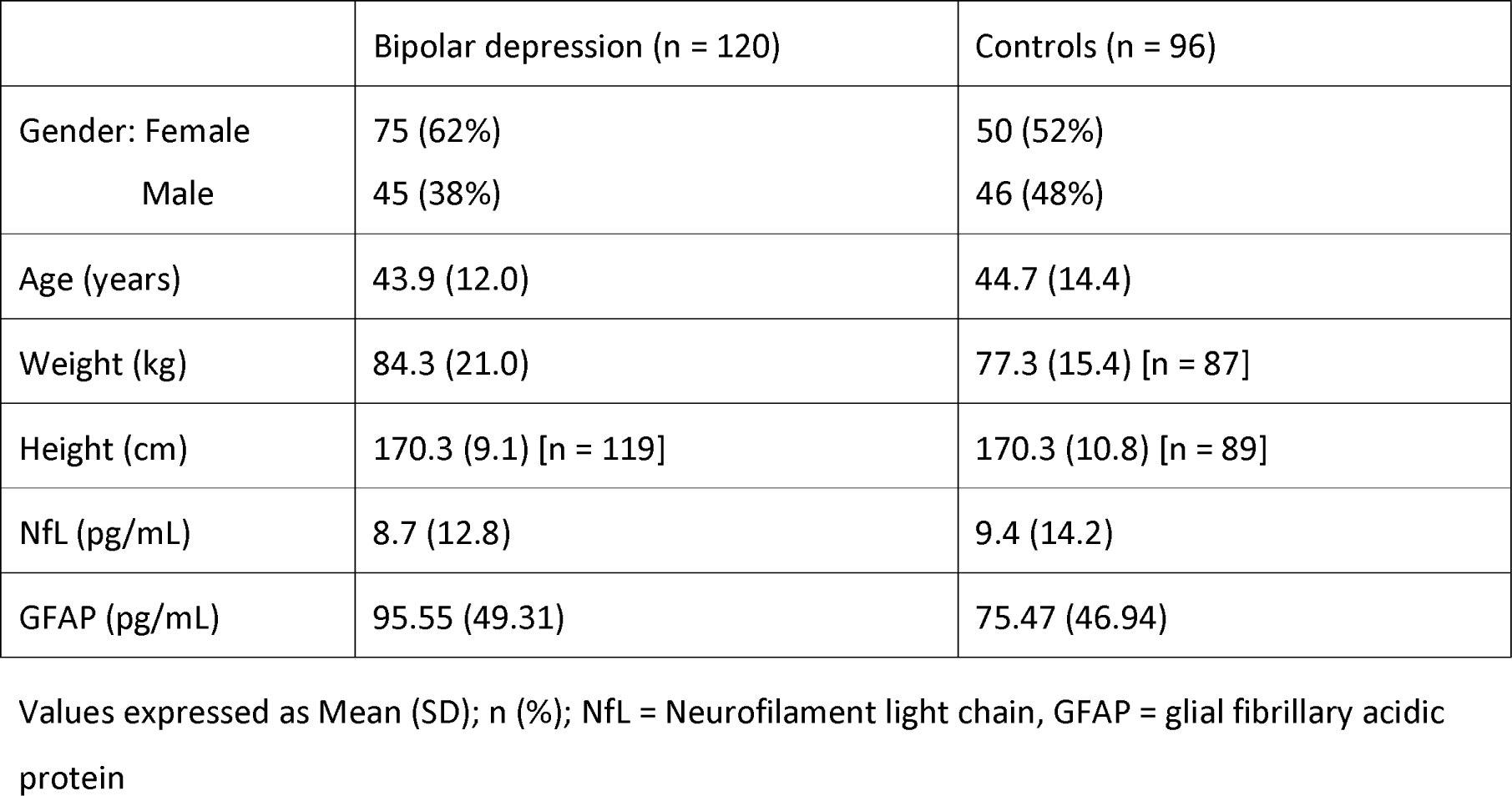
Descriptive comparison of the cohorts.

|  | Bipolar depression (n = 120) | Controls (n = 96) |
| --- | --- | --- |
| Gender: Female | 75 (62%) | 50 (52%) |
| Male | 45 (38%) | 46 (48%) |
| Age (years) | 43.9 (12.0) | 44.7 (14.4) |
| Weight (kg) | 84.3 (21.0) | 77.3 (15.4) [n = 87] |
| Height (cm) | 170.3 (9.1) [n = 119] | 170.3 (10.8) [n = 89] |
| NfL (pg/mL) | 8.7 (12.8) | 9.4 (14.2) |
| GFAP (pg/mL) | 95.55 (49.31) | 75.47 (46.94) |

Values expressed as Mean (SD); n (%); NfL = Neurofilament light chain, GFAP = glial fibrillary acidic protein

As for the clinical characteristics of the bipolar depression cohort, their mean age at diagnosis was 34 ± 11 years old, duration of bipolar illness was 23 ± 11 years, 78 (65%) individuals were prescribed at least one mood stabiliser, and 75 (69%) individuals reported having experienced 20+ depressive episodes in their lifetime. With regards to their symptom severity and functioning, their mean MADRS was 29.2 ± 5.2, BDRS was 25.7 ± 6.6, HAM-A was 17.6 ± 5.7, YMRS was 3.9 ± 3.4, Q-LES-Q was 40.1 ± 12.7 and CGI-S was 4.7 ± 0.8.

### Biomarker levels in bipolar depression and healthy controls

The raw mean ± SD of plasma NfL was 8.7 ± 12.8pg/mL in bipolar depression and 9.4 ± 14.2pg/mL in healthy controls. The raw mean ± SD of GFAP was 95.55 ± 49.31 in bipolar depression and 75.47 ± 46.94 in healthy controls. Plasma GFAP was significantly elevated in people with bipolar depression compared to healthy controls after adjusting for age, sex and weight (β=0.21 [0.07, 0.35], p=0.006). Consistent with our previous finding,^13^ log-transformed NfL was also significantly elevated in people with bipolar depression compared to healthy controls (β=0.06 [0.01, 0.10], p=0.028).

### Clinical correlates of NfL and GFAP

Age positively correlated with both NfL and GFAP (**Table 2**). People who had a history of substance use disorder (SUD) in their lifetime had significantly lower GFAP concentration than those without (t = 2.970, df = 95.753, adjusted p = 0.023). However, people with a SUD history were younger than those without (mean 38.15 ± 8.63 years vs 46.44 ± 12.38 years; F = 13.63, p < 0.001). The multivariable GLM including age and SUD history found that SUD was not significantly associated with GFAP (B = −0.33; [-0.68, 0.02]; p = 0.058), suggesting therefore, that the difference was attributable to age.

**Table 2-.**
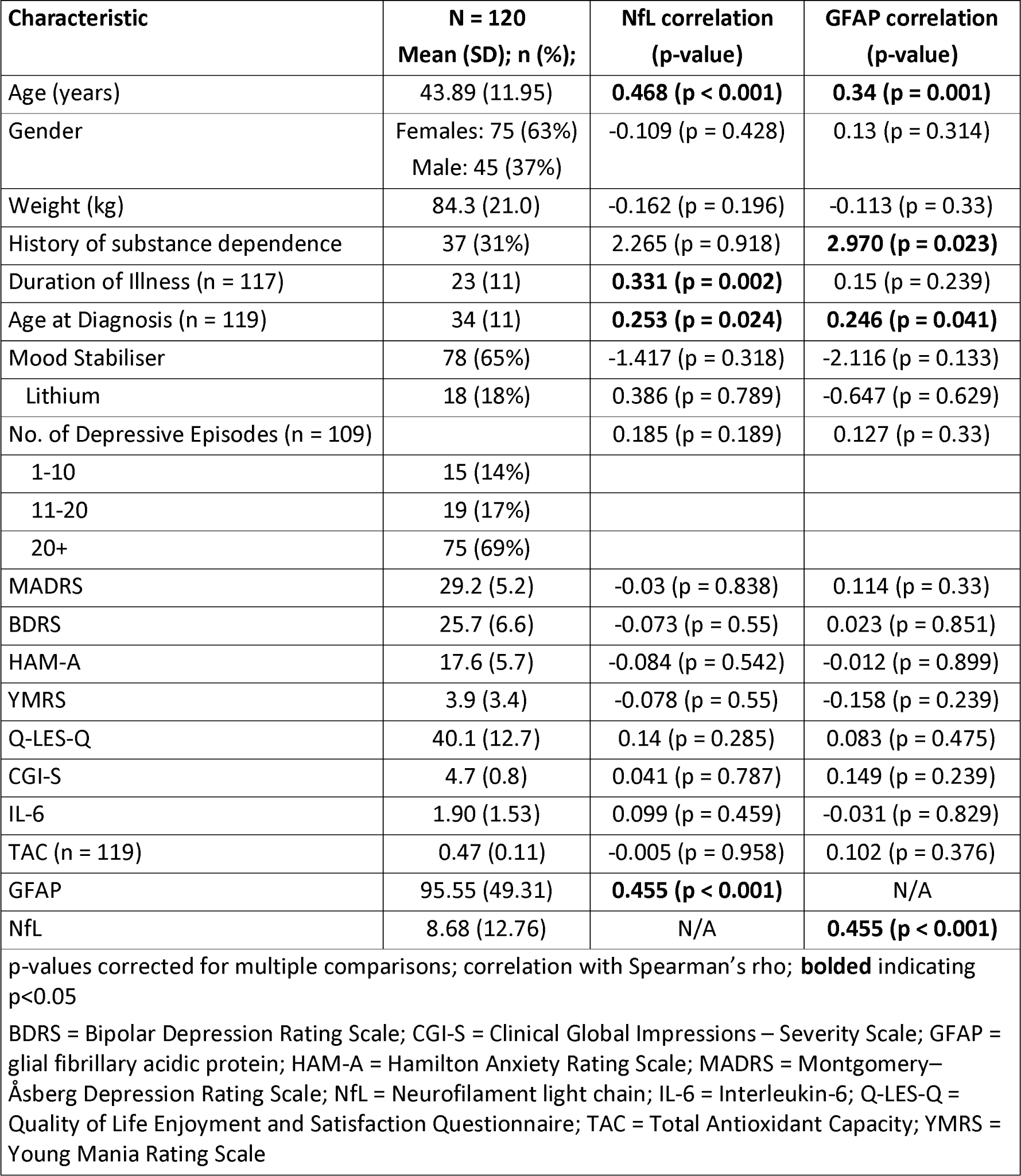
Clinical correlates of NfL and GFAP in bipolar depression.

Both duration of illness and age at diagnosis significantly correlated with NfL concentration (Supplementary Figure 1), while only age at diagnosis correlated with GFAP concentration (Supplementary Figure 2). Duration of illness (VIF = 1.69) and age at diagnosis (VIF = 2.08) were not significant for multicollinearity (VIF < 3.0). Neither duration of illness nor age at diagnosis were significant in the GLMs when they were added with age to predict NfL or GFAP.

### BMA analysis

#### Plasma NfL

BMA revealed several important predictors of NfL. The posterior inclusion probabilities (PIP) are shown in Figure 1a. The strongest evidence was observed for GFAP (PIP = 1), whilst duration of illness was also strongly supported (PIP = 0.85). Weight (PIP = 0.50) was supported as a strong predictor in the BMA analysis without number of depressive episodes (n = 120), but not in the smaller BMA model (n = 109) which included the number of depressive episodes (PIP = 0.28). The remaining predictors (Table 3), including age, had PIPs < 0.5, and only GFAP was a significant independent predictor of NfL, with its 95% CI not capturing zero. Taken in the context of the PIP value > 0.5, this indicates that GFAP is important to include in any well-fitting model predicting NfL using the possible variables, whilst there is some doubt about the magnitude of the other predictors in predicting NfL. A visual representation of the importance of each predictor in the top 20 models is included in the supplementary material (sFigure 3 and sFIgure 4).

**Figure 1a-.**
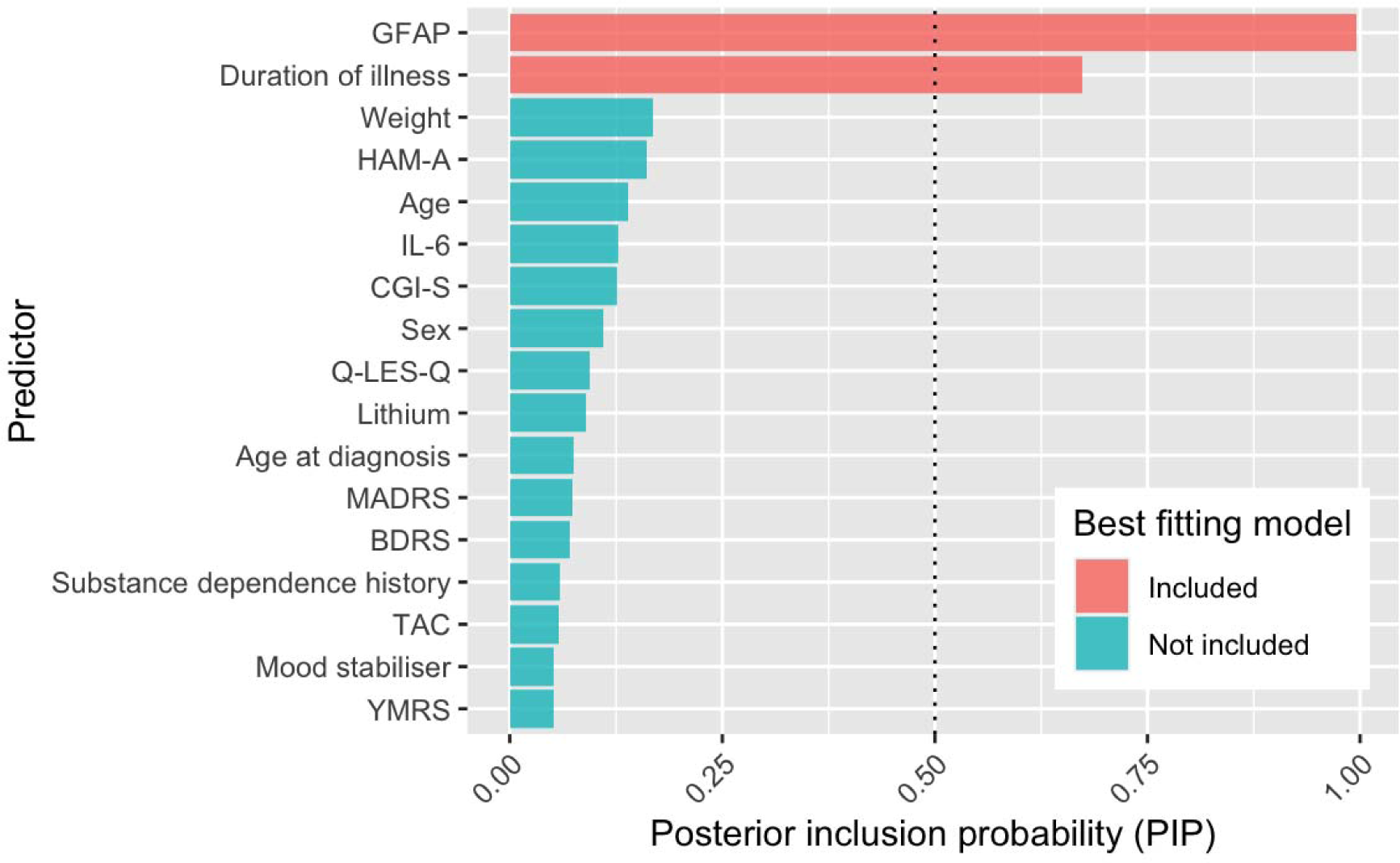
PIP for each predictor of NfL and model parameters averaged across the model space.

**Table 3=.** BMA analysis for prediction of NfL concentration.

| Predictor | B [95% CI] | PIP |
| --- | --- | --- |
| Age | 0.03 [0 - 0.16] | 0.29 |
| Sex | -0.13 [-1.63 – 0] | 0.14 |
| Weight | -0.02 [-0.07 - 0] | 0.5 |
| Substance dependence history | -0.04 [-1.06 - 0.01] | 0.09 |
| Duration of illness | 0.12 [0 - 0.19] | 0.85 |
| Age at diagnosis | 0 [-0.05 - 0.02] | 0.1 |
| Mood stabiliser | 0.03 [-0.09 -0.79] | 0.09 |
| Lithium | -0.11 [-1.59 -0.05] | 0.12 |
| MADRS | 0 [-0.11 - 0.02] | 0.1 |
| BDRS | 0 [-0.08 - 0.03] | 0.1 |
| HAM-A | -0.05 [-0.21 - 0] | 0.34 |
| YMRS | 0 [-0.08 - 0.06] | 0.08 |
| Q-LES-Q | 0 [0 - 0.06] | 0.12 |
| CGI-S | -0.11 [-1.16 – 0] | 0.16 |
| IL-6 | 0.14 [0 - 0.77] | 0.28 |
| TAC | -0.13 [-4.02 - 0.26] | 0.09 |
| GFAP | 0.04 [0.02 - 0.05] | 1 |

B = model coefficients computed as the mean of the posterior distribution; SE = standard deviation of the posterior distributions; 95% CI = 95% credible intervals. PIP = posterior inclusion probability

#### Plasma GFAP

NfL (PIP = 1) was considered an important predictor (Figure 1b and Table 4), whilst age at diagnosis was a moderately strong predictor (PIP = 0.67) but its estimate was less precise and included 0 in its 95% CI (B = 0.78, 95% CI: 0 – 1.74), which taken together implies that this predictor is important to include in any well-fitting model, but there is considerable uncertainty regarding the specific magnitude of this relationship. The other variables, including age, were not considered to be strong predictors of GFAP.

**Figure 1b-.**
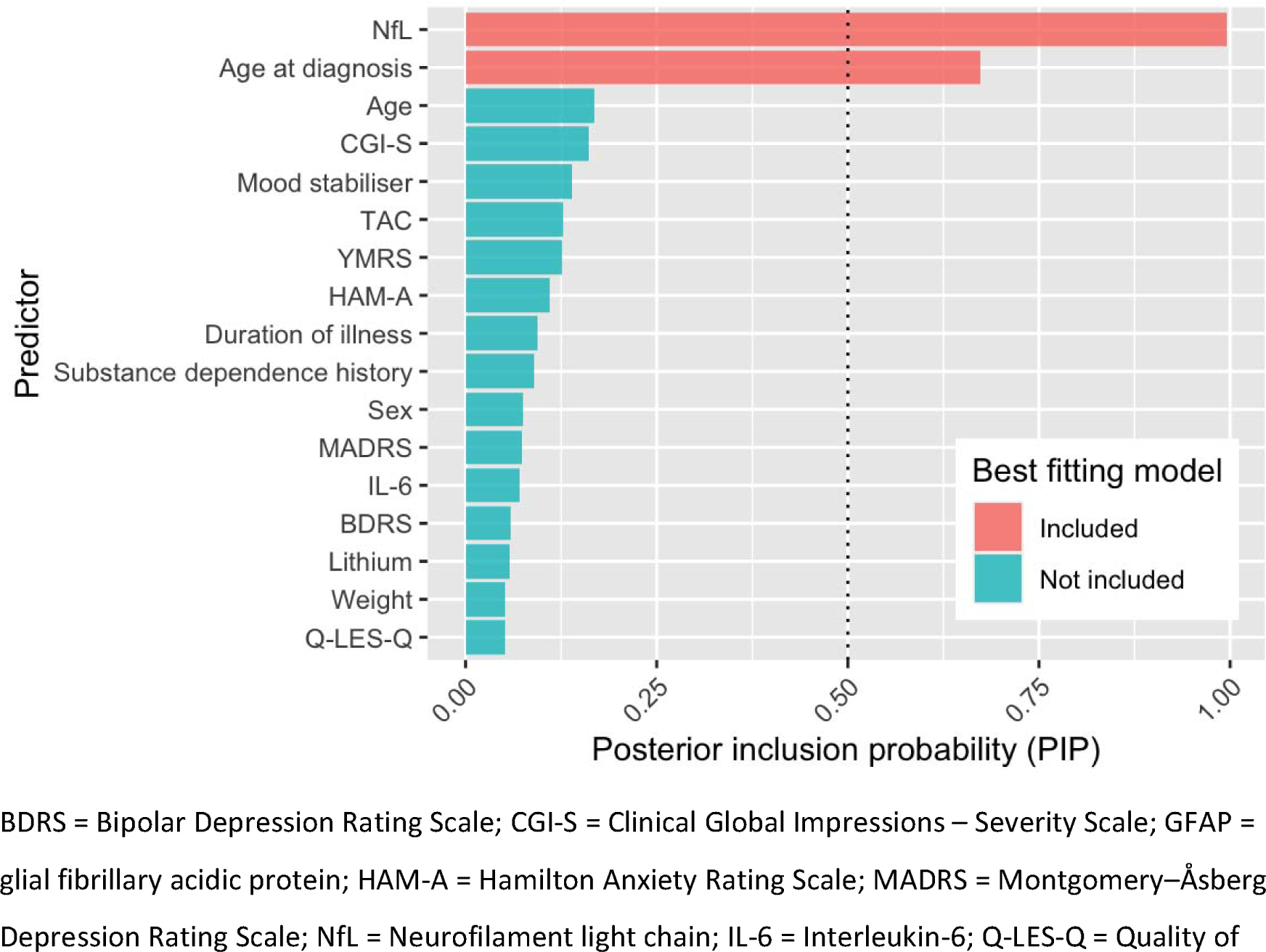
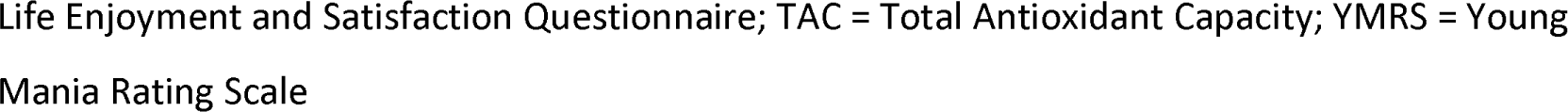
PIP for each predictor of GFAP and model parameters averaged across the model space.

**Table 4=.** BMA analysis for prediction of GFAP concentration.

| Predictor | B [95% CI] | PIP |
| --- | --- | --- |
| intercept | 96.28 [88.07 - 104.16] | 1 |
| Age | 0.14 [0 – 1.32] | 0.17 |
| Sex | 0.61 [0 – 7.5] | 0.08 |
| Weight | 0 [-0.02 – 0] | 0.05 |
| Substance dependence history | -0.97 [-13.8 – 0] | 0.09 |
| Duration of illness | -0.06 [-0.71 – 0] | 0.09 |
| Age at diagnosis | 0.78 [0 – 1.74] | 0.67 |
| Mood stabiliser | 1.84 [0 – 18.96] | 0.14 |
| Lithium | 0.35 [-0.33 – 3.3] | 0.06 |
| MADRS | 0.04 [-0.01 – 0.76] | 0.07 |
| BDRS | 0 [-0.16 – 0.01] | 0.06 |
| HAM-A | 0.11 [0 – 1.31] | 0.11 |
| YMRS | -0.25 [-2.66 – 0.04] | 0.13 |
| Q-LES-Q | 0 [-0.03 – 0] | 0.05 |
| CGI-S | 1.46 [0 – 12.45] | 0.16 |
| IL-6 | -0.17 [-2.61 – 0] | 0.07 |
| TAC | 6.77 [0 – 70.66] | 0.13 |
| NfL | 4.13 [2.17 – 6.03] | 1 |

B = model coefficients computed as the mean of the posterior distribution; SE = standard deviation of the posterior distributions; 95% CI = 95% credible intervals. B = odds ratios of the model coefficients; PIP = posterior inclusion probability

## DISCUSSION

This is the largest study to date exploring the relationship between brain-specific markers of neurodegeneration and astroglial dysfunction in people with bipolar depression. By extending on our previous work,^13^ we found that both plasma NfL and GFAP are elevated in people with bipolar depression. Plasma NfL was associated with a longer duration of illness and may represent a marker of illness stage. Our novel finding that plasma GFAP was elevated in bipolar depression suggests that there is increased activation of astrocytes, which has been linked with neuroinflammation, adding to the debated literature about the role of inflammation in bipolar disorder.^24–26^ Plasma GFAP was also positively associated with a later age of bipolar diagnosis, which suggests that those with a later age of onset have a distinct neurobiological process.

The difference in NfL concentrations between people with bipolar disorder and unaffected individuals was small and only evident after adjusting for known covariates. This contrasts with the markedly elevated concentrations (at least two-fold) seen in neurodegenerative disorders such as dementia and multiple sclerosis,^27,28^ suggesting that the subtle increase in bipolar disorder is due to a different process. Aggio et al’s study^12^ similarly found that NfL was mildly elevated in people with bipolar depression (n=45) and linked this to neuronal distress with axonal damage due to an ageing-related process, or “accelerated brain ageing”. Accelerated brain ageing refers to the age gap between chronological and the estimated neuroanatomical age, which has been observed in various psychiatric disorders including bipolar disorder.^29,30^ Plasma NfL has been proposed as a potential marker of brain ageing^31,32^ due to its age-related increases.^33^ As the brain atrophies with age,^34^ NfL is released from dendrites and axons into CSF and blood.^35^ The subtle elevation of plasma NfL levels in bipolar disorder after adjustment for chronological age supports the notion that there is accelerated ageing in bipolar disorder, adding to the current debated literature.^29,30,36^

Moreover, our finding that plasma NfL levels correlate with the duration of bipolar disorder may be evidence of the progressive burden of the condition on the brain. This supports the “neuroprogression” hypothesis, which postulates that a subset of people have a progressive trajectory due to pathological rewiring and degeneration of the brain over the course of the illness.^9,37^ Longitudinal neuroimaging studies have found that people with bipolar disorder have faster cortical atrophy^38^ and ventricular enlargement^39^ beyond what is expected with normal aging.^40^ The BMA analysis further supported the notion that duration of bipolar disorder was an important predictor of plasma NfL, or brain age.

The BMA analysis also identified that duration of illness, and not chronological age, was more important in predicting plasma NfL. This negative finding of chorological age not predicting plasma NfL was surprising given it is contrary to well-established literature,^33^ including our own conventional frequentist analysis in this study. This contradiction highlights the strength of BMA analysis in being able to consider a broad range of possible predictors, examine a wide range of models based on all the variables available, and provide a value that represents the importance of each predictor across all possible models. This contrasts with the traditional frequentist approach, which can only consider one model at a time introducing bias.^41^ Given that plasma NfL level is impacted by many factors,^5^ not limited to the person’s age, weight, and physical health, the ability of BMA to consider a wide range of predictors uncovered an interesting insight in this study. Nonetheless, the limited age range of our cohort warrants further research incorporating younger and older cohorts with bipolar disorder. In addition, studies investigating the three-way relationship between NfL, neuroimaging, and the burden of bipolar disorder (longer duration, number of manic and/or depressive episodes) is required to validate this hypothesis that the duration of bipolar illness has a greater burden on the brain age than chronological age.

Our finding of elevated GFAP levels in bipolar disorder suggests that neuroinflammation may be a component of this condition, especially in those with later onset of illness. This may be connected to the increased prevalence of cardiovascular and metabolic conditions in older adults, which are known to cause neuroinflammatory changes and oxidative stress in the brain.^42^ Although we did not collect details about their cardiovascular health, the Clinical and Health Outcomes Initiative in Comparative Effectiveness for Bipolar Disorder study (Bipolar CHOICE), which had a comparable cohort age (38.9 ± 12.1 years), found that individuals with a later age onset of bipolar disorder were more likely to have underlying medical comorbidities.^43^ Combining this with our finding that GFAP was higher in those with later age of onset, this may represent an increased expression and release of GFAP from astrocytic activation as a form of defense response to neuroinflammation. However, GFAP did not correlate with peripheral interleukin-6 (IL-6) or total antioxidant capacity (TAC), suggesting that peripheral and CNS inflammation may contribute to the pathophysiology of bipolar disorder in distinct ways. Future studies should characterise cardiovascular and metabolic risk and compare this with biomarker levels.

It remains unclear whether mild increases in plasma NfL and/or GFAP in bipolar disorder are related to the trait (bipolar disorder) or state (depression, mania or mixed state).^44^ Previous research by Steinacker et al. found that GFAP correlated with the severity of unipolar depressed state as measured by the MADRS.^7^ We found that the severity of depressive symptoms did not correlate with plasma NfL nor GFAP suggests that unipolar and bipolar depression may have different underlying neurobiological processes. Despite lithium’s postulated neuroprotective role,^45^ it also did not have a significant effect on plasma NfL nor GFAP, though it may have been underpowered considering that plasma NfL and GFAP levels were only mildly elevated in people with bipolar disorder.

This study has several limitations. The cross-sectional nature of the study makes it difficult to make causal inferences, so future studies with serial samples and longitudinal clinical data are needed. We lacked other important clinical information about the people with bipolar disorder, including cognition and number of manic episodes. Investigating these biomarkers in manic states may yield important insights, although recruiting participants in acute mania is challenging. Furthermore, detailed history including quantification of substance use would have been useful to better understand the correlation between substance use disorder history and GFAP, highlighting a potential area for further research. Finally, we recommend future research to use a multimodal approach, combining fluid-based biomarker levels with neuroimaging to strengthen the findings.

In conclusion, this is the largest study to date that has investigated plasma NfL and GFAP in people with bipolar disorder. We found that both are mildly elevated compared to healthy controls, with higher NfL associated with a longer duration of bipolar disorder, while GFAP was associated with a later age of onset. Taken together, our study further supports the multimodal theory of bipolar disorder, which includes neuroinflammatory and axonal pathology components. Moreover, these biomarkers may be potential candidates to identify different stages and phenotypes of bipolar disorder, such as neuroprogression or later onset of illness. Further studies with longitudinal data are needed to further validate our findings and better understand the neurobiological underpinnings of bipolar disorder.

## Data Availability

Deidentified and pooled data is available at request to the corresponding author.

## CRediT authorship contribution statement

Matthew Kang: Study design, data analysis and original manuscript writing

Dhamidhu Eratne: Study design, data analysis, manuscript revision and supervision

Charles Malpas: Study design, data analysis, statistical support and supervision

Alexander F Santillo: Study design, data analysis, statistical support and supervision

Claudia Cicognola: Manuscript revision

Shorena Janelidze: Data analysis, manuscription revision

Olivia Dean: Study design, manuscript revision

Oskar Hansson: Study design, manuscript revision

Michael Berk: Study design, manuscript revision

Adam J Walker: Study design, manuscript revision

Jasleen Grewal: Manuscript revision

Dennis Velakoulis: Study design, data analysis, manuscript revision and supervision

Philip B Mitchell: Manuscript revision and supervision

Christos Pantelis: Study design, Manuscript revision and supervision

Malcolm Hopwood: Manuscript revision and supervision

## ACKNOWLEDGEMENTS AND FUNDING SOURCES

This study was supported by: MACH MRFF RART 2.2, NHMRC (1185180), and Psychiatry and Rehabilitation Division, Region Skåne, Sweden. The role of these funding sources was to support research study staff and biosample analyses. The healthy controls cohort was part of a major Australian Department of Industry Co-operative Research Centre (CRC) grant - https://researchdata.ands.org.au/treatment-resistant-schizophrenia-biobank/1325206

MK is supported by the Nick Christopher PhD scholarship and the Research Training Program Scholarship from the Department of Psychiatry, University of Melbourne with contributions from the Australian Commonwealth Government, and the Ramsay Hospital Research Foundation.

AFS is supported by the Swedish federal government under the ALF agreement.

MB is supported by a NHMRC Leadership 3 Investigator grant (GNT2017131).

C Pantelis was supported by a National Health and Medical Research Council (NHMRC) Senior Principal Research Fellowship (1105825), an NHMRC L3 Investigator Grant (1196508).

PBM is supported by an NHMRC Investigator Grant (1177991). In the last 3 years he has received remuneration from Janssen (Australia) for lectures and advisory board membership

OH’s work at Lund University was supported by Alzheimer’s Association (ZEN24-1069572, SG-23-1061717), GHR Foundation, Swedish Research Council (2022-00775), ERA PerMed (ERAPERMED2021-184), Knut and Alice Wallenberg foundation (2022-0231), Strategic Research Area MultiPark (Multidisciplinary Research in Parkinson’s disease) at Lund University, Swedish Alzheimer Foundation (AF-980907), Swedish Brain Foundation (FO2021-0293), Parkinson foundation of Sweden (1412/22), Cure Alzheimer’s fund, Rönström Family Foundation, Konung Gustaf V:s och Drottning Victorias Frimurarestiftelse, Skåne University Hospital Foundation (2020-O000028), Regionalt Forskningsstöd (2022-1259) and Swedish federal government under the ALF agreement (2022-Projekt0080).

AJW was previously supported by a Trisno Family Research Fellowship.

The corresponding author had full access to all the data in the study and had final responsibility for the decision to submit for publication.

## DATA SHARING STATEMENT

Deidentified and pooled data is available at request to the corresponding author.

