## Supplemental figures for "Plasma Glial Fibrillary Acidic Protein and Neurofilament Light are Elevated in Bipolar Disorder: Evidence for Neuroprogression and Astrocytic Activation"

Figure s1 – Scatterplot of plasma NfL and duration of illnses
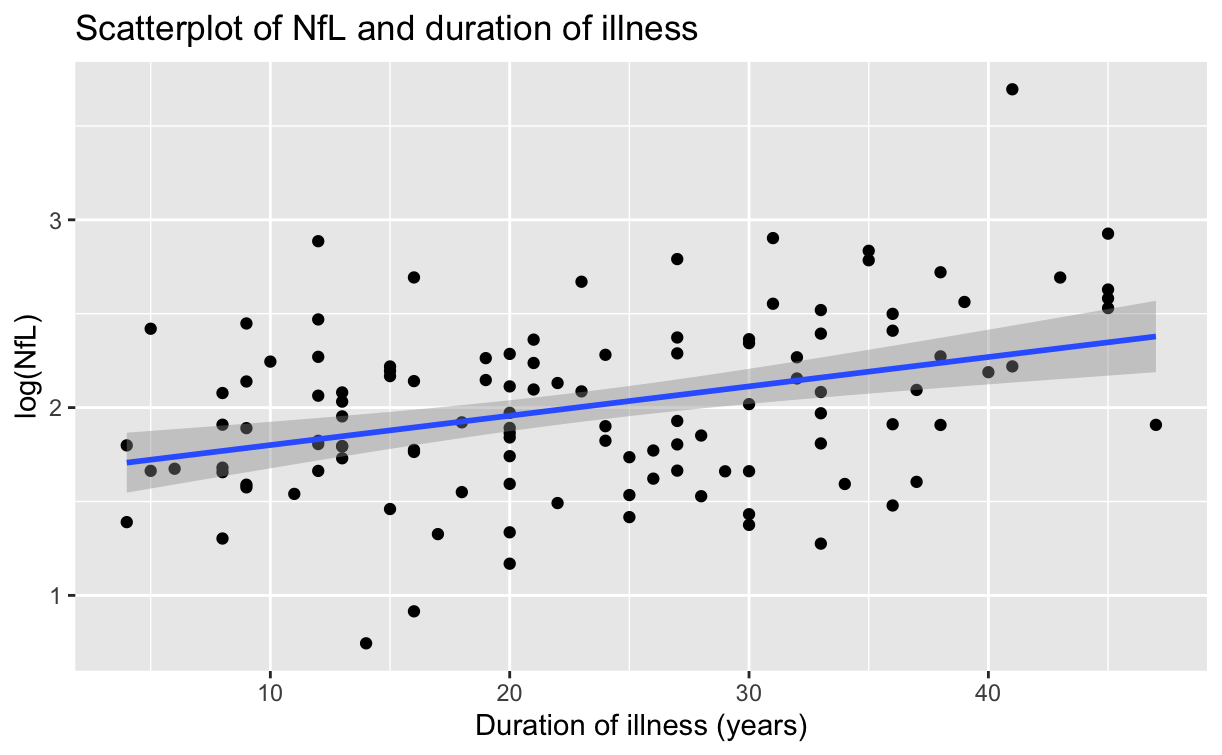

Figure s2 – Scatterplot of plasma GFAP and age at diagnosis
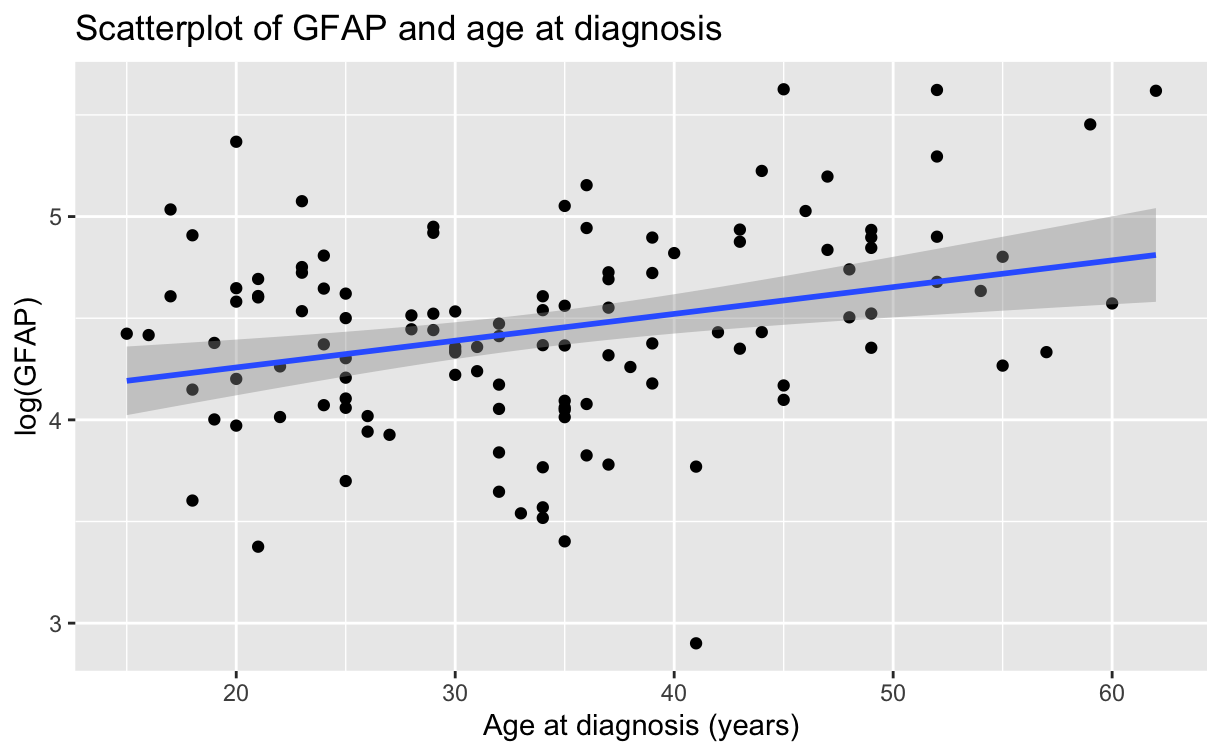

Figure s3 - Visualisation of the model space from the BMA analysis of NfL correlates

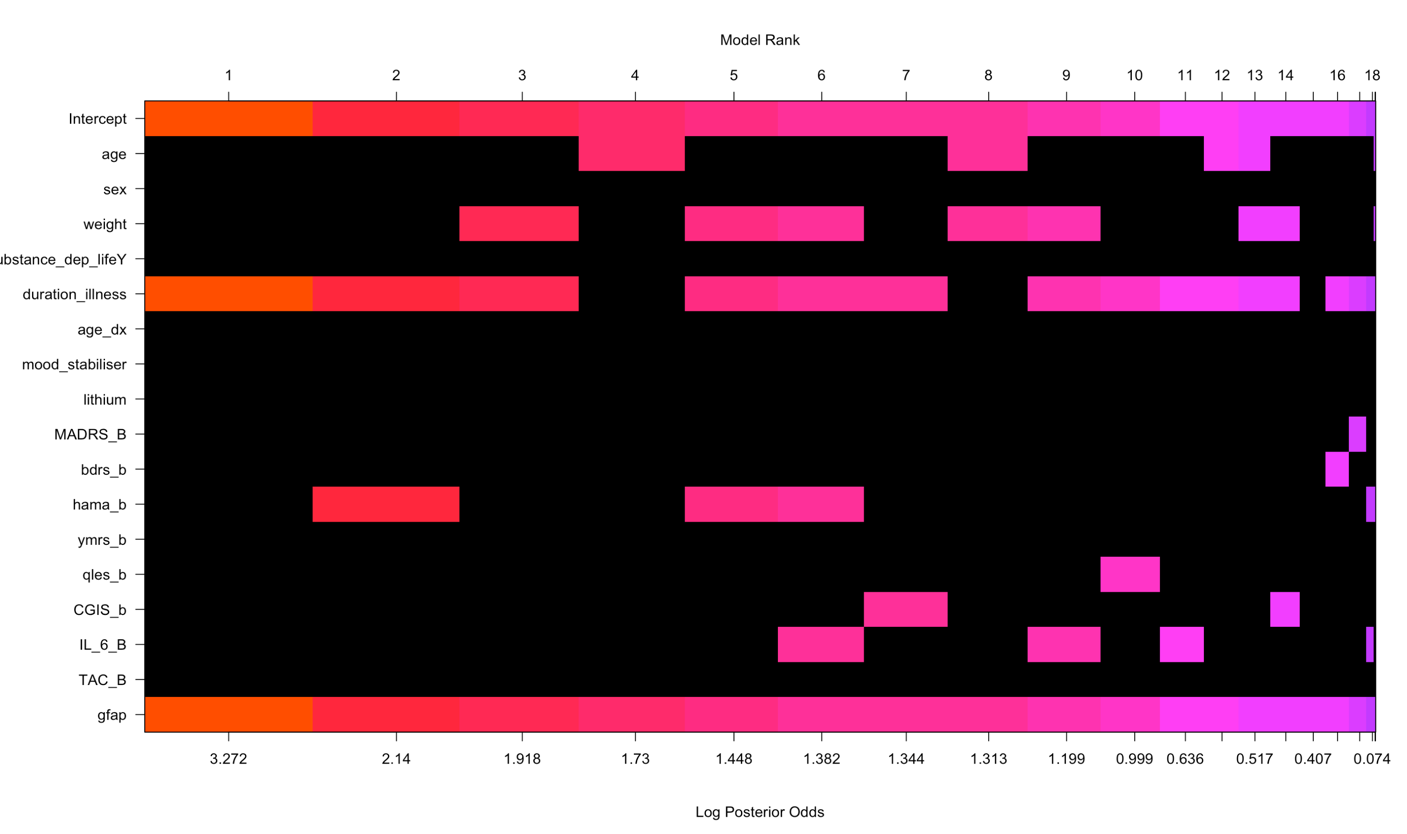

BDRS = Bipolar Depression Rating Scale; CGI-S = Clinical Global Impressions – Severity Scale; GFAP = glial fibrillary acidic protein; HAM-A = Hamilton Anxiety Rating Scale; MADRS = Montgomery–Åsberg Depression Rating Scale; NfL = Neurofilament light chain; IL-6 = Interleukin-6; Q-LES-Q = Quality of Life Enjoyment and Satisfaction Questionnaire; TAC = Total Antioxidant Capacity; YMRS = Young Mania Rating Scale

Figure s4 - Visualisation of the model space from the BMA analysis of GFAP correlates

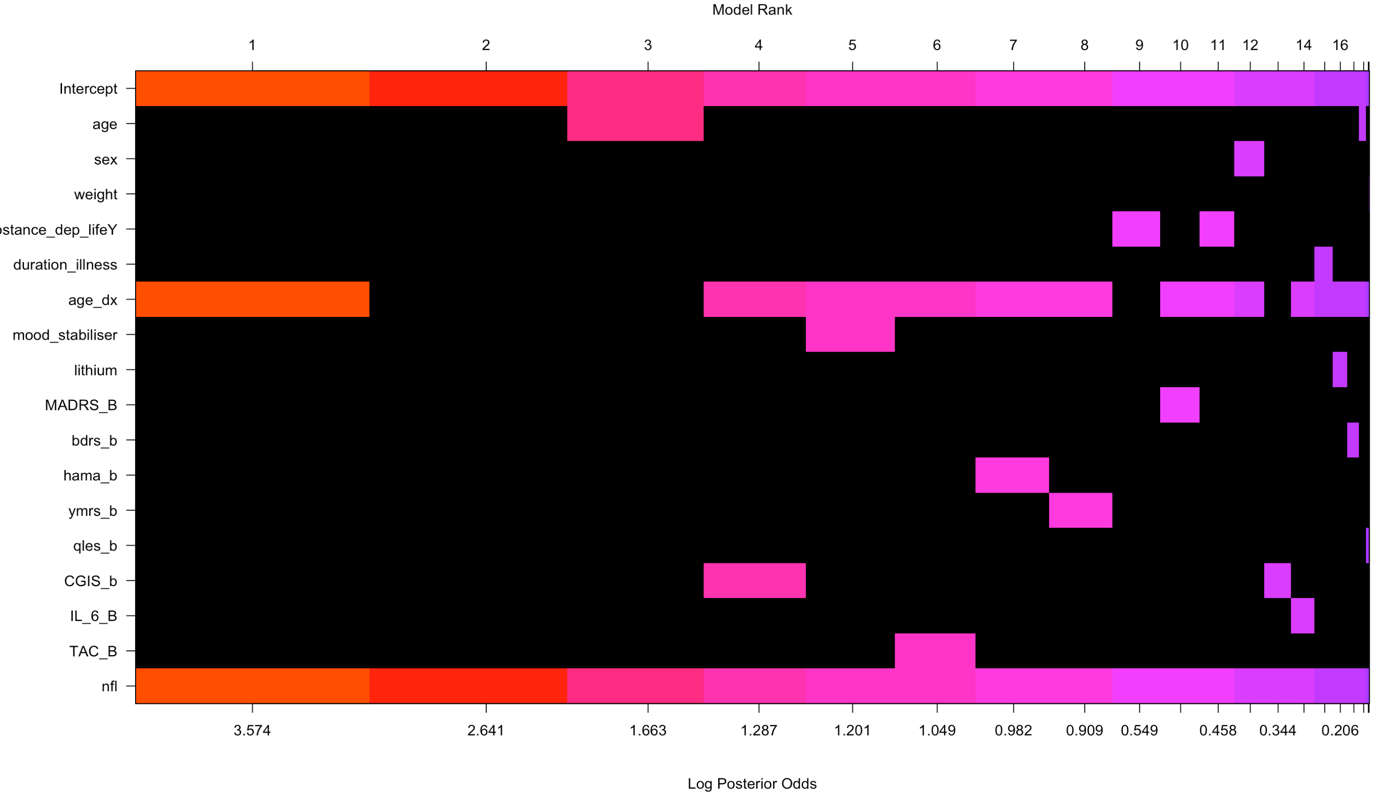

BDRS = Bipolar Depression Rating Scale; CGI-S = Clinical Global Impressions – Severity Scale; GFAP = glial fibrillary acidic protein; HAM-A = Hamilton Anxiety Rating Scale; MADRS = Montgomery–Åsberg Depression Rating Scale; NfL = Neurofilament light chain; IL-6 = Interleukin-6; Q-LES-Q = Quality of Life Enjoyment and Satisfaction Questionnaire; TAC = Total Antioxidant Capacity; YMRS = Young Mania Rating Scale
